# Association Between SARS-CoV-2 RNAemia and Post-Acute Sequelae of COVID-19

**DOI:** 10.1101/2021.09.03.21262934

**Authors:** Nikhil Ram-Mohan, David Kim, Angela J Rogers, Catherine A Blish, Kari C Nadeau, Andra L Blomkalns, Samuel Yang

## Abstract

Determinants of Post-Acute Sequelae of COVID-19 are not known. Here we show that 75% of patients with viral RNA in blood (RNAemia) at presentation were symptomatic in the post-acute phase. RNAemia at presentation successfully predicted PASC, independent of patient demographics, initial disease severity, and length of symptoms.

## Background

The determinants of COVID-19 severity and extrapulmonary complications have now been well studied, and RNAemia (viral RNA in blood) has emerged as an important factor (1,2). Much less is known about the determinants of Post-Acute Sequelae of COVID-19 (PASC), the persistence or development of new symptoms after the acute phase of infection, recently reported to affect as many as 87.4% of COVID-19 patients (3,4) primarily with moderate or worse severity (5,6). Recent evidence suggested persistent clotting protein pathology with elevated levels of antiplasmin (7) and non-classical monocytes (8) in patients with PASC. Discovery of SARS-CoV-2 S1 protein in these non-classical monocytes and fragmented SARS-CoV-2 RNA in peripheral blood mononuclear cells in a PASC patient 15 months post infection further exhibited the persistence of viral particles (8). Given the importance of RNAemia in disease severity and its persistence in the blood, we describe the relationship between RNAemia at presentation and post-acute symptoms at least three weeks after symptom onset.

## Methods

We studied the clinical trajectories of 155 patients enrolled in the IRB-approved (eP-55650) Stanford Hospital Emergency Department (ED) COVID-19 Biobank between April and November 2020 with completed follow-ups. We assessed symptoms and severity (based on a modified WHO scale) (1) on the date of enrollment (median = 4, range = 0 – 44 days after symptom onset), and at least three weeks after symptom onset (median = 35, range = 21 – 79 days).

We measured SARS-CoV-2 RNAemia at the time of enrollment, using the definitions of our earlier study (1). We compared the proportions of initially RNAemic and non-RNAemic patients with persistent or new symptoms in the post-acute phase using a 2-sample chi-squared test with continuity correction. We estimated the association between RNAemia at enrollment and PASC at follow-up in a logistic model controlling for disease severity at enrollment, patient demographics (age and gender), presence of any symptom at enrollment (anxiety, dizziness, fatigue, hair loss, palpitations, rash, insomnia, chest pain, chills, cough, decrease in sense of taste, fever, nausea/vomiting/diarrhea, headache, loss of smell, myalgia, new confusion, shortness of breath), and durations of symptoms. We also compared the median number of PASC symptoms for RNAemic and non-RNAemic patients using the Wilcoxon rank-sum test with continuity correction. We performed all analyses in R (version 4.0.3).

## Results

49.0% (76/155) of patients were women, and the median age was 45 years (IQR 34 – 60). At enrollment, 27.1% (42/155) of patients had mild disease severity, 67.7% (105/155) moderate, and 5.2% (8/155) severe. Patients had a median of six symptoms (IQR = 4–8): 72.3% (112/155) had a cough, 65.3% (101/155) had shortness of breath, and 64.5% (99/155) had fever. In the post-acute phase, 52.3% (81/155) had one or more new (24.5% [38/155]) or persistent (37.4% [58/155]) symptoms, of which the most common were cough, dizziness, and loss of smell (Table 1). 1.3% (2/155) of patients developed anxiety that was not present at enrollment.

**Table:** Progression of COVID-19 symptoms.

| Symptom | On enrollment | Persistent at follow-up | New at follow-up |
| --- | --- | --- | --- |
| **Cardiovascular** |  |  |  |
| Chest Pain | 34.2% (53/155) | 11.3% (6/53) | 0.7% (1/149) |
| Palpitations | 0.6% (1/155) | 0 (0) | 1.3% (2/155) |
| **Dermatologic** |  |  |  |
| Rash | 1.3% (2/155) | 0 (0) | 0.6% (1/155) |
| Hair Loss | 0 (0) | 0 (0) | 0.6% (1/155) |
| **Gastrointestinal** |  |  |  |
| Nausea/vomiting/diarrhea | 43.8% (68/155) | 7.3% (5/68) | 0.7% (1/150) |
| **Constitutional** |  |  |  |
| Fever | 64.5% (100/155) | 1.0% (1/100) | 0 (0) |
| Chills | 32.9% (51/155) | 3.9% (2/51) | 0.6% (1/153) |
| Myalgia | 41.2% (64/155) | 15.6% (10/64) | 4.1% (6/145) |
| Fatigue | 36.7% (57/155) | 14.0% (8/57) | 8.2% (12/147) |
| **Neuropsychiatric** |  |  |  |
| Loss of Taste | 41.2% (64/155) | 14.0% (9/64) | 0 (0) |
| Loss of Smell | 29.6% (46/155) | 19.5% (9/46) | 0 (0) |
| Confusion | 2.5% (4/155) | 0 (0) | 2.6% (4/155) |
| Headache | 22.5% (35/155) | 2.8% (1/35) | 5.2% (8/154) |
| Dizziness | 5.8% (9/155) | 22.2% (2/9) | 2.6% (4/153) |
| Insomnia | 0.6% (1/155) | 0 (0) | 0.6% (1/155) |
| Anxiety | 0 (0) | 0 (0) | 1.3% (2/155) |
| **Respiratory** |  |  |  |
| Cough | 72.2% (112/155) | 25.8% (29/112) | 3.9% (5/126) |
| Shortness of breath | 65.1% (101/155) | 18.8% (19/101) | 3.7% (5/136) |

75.0% (27/36) of initially RNAemic patients were symptomatic in the post-acute phase, compared to 43.7% (52/119) of non-RNAemic patients (difference = 31.3% [95% CI, 12.8% - 49.8%], p=0.002). RNAemic patients had a median of one symptom in the post-acute phase compared to zero in non-RNAemic patients (p=0.014, Wilcoxon rank-sum test). RNAemia at presentation predicted PASC, conditional on patient demographics and initial disease severity (OR 1.31 [95% CI, 1.08 – 1.59], p=0.007 (Supplement)). The association was strongest for patients with moderate disease severity at presentation (Figure), with 78.6% (22/28) of initially RNAemic patients symptomatic in the post-acute phase, compared to 45.5% (35/77) of non-RNAemic patients (difference = 33.1% [95% CI, 11.8% - 54.4%], p=0.005). This difference was due almost entirely to persistent or new respiratory symptoms (difference in proportions = 28.2% [95% CI, 8.4% - 47.9%], p=0.002).

**Figure.**
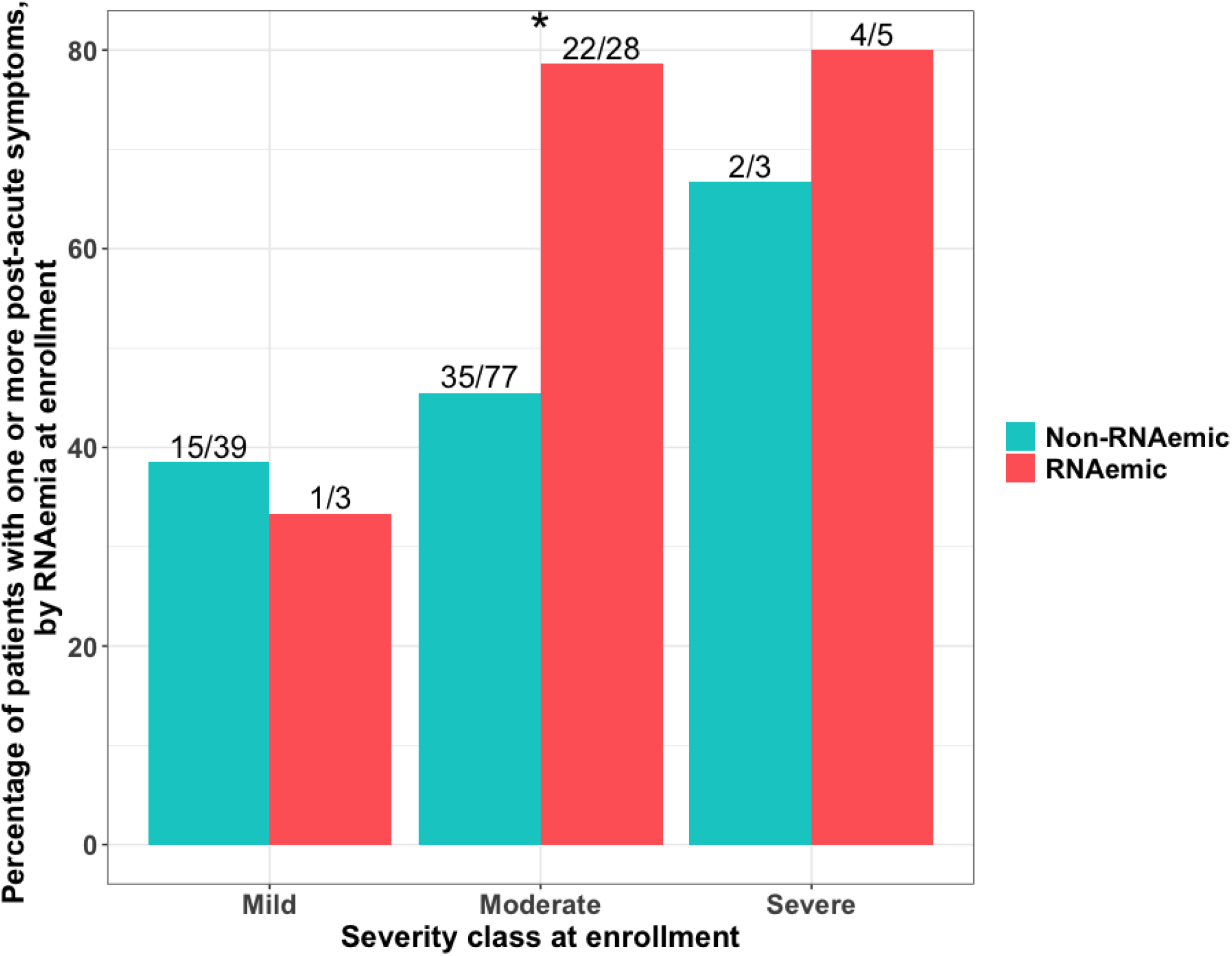
Rate of post-acute sequelae of SARS-CoV-2 infection, by RNAemia and clinical severity on enrollment. Overall, 75.0% (27/36) of initially RNAemic patients had one or more post-acute symptoms at follow-up, compared to 43.7% (52/119) of non-RNAemic patients (difference = 31.3% [95% CI: 12.8% - 49.8%], p=0.002). Conditional on severity at enrollment (mild = discharged from ED [n=42], moderate = hospitalized, requiring no more than oxygen by nasal cannula [n=105], severe = hospitalized, requiring high-flow nasal cannula or mechanical ventilation [n=8]), RNAemia on presentation was associated with significantly higher rates of PASC for presentations of moderate severity (difference = 33.1% [95% CI, 11.8% - 54.4%], p=0.005). * indicates p-value<0.05.

## Discussion

To our knowledge, this study describes the first reported association between SARS-CoV-2 RNAemia and PASC. RNAemia at presentation was associated with new or persistent symptoms at least 21 days after symptom onset independent of initial patient severity and the association was strongest among patients with moderately severe clinical presentations requiring hospital admission. This finding adds to the growing literature on SARS-CoV-2 RNAemia’s role in disease severity and extrapulmonary complications in the acute phase of illness, as well as the association between hospitalization and PASC (1,2,5,6). The incidence of PASC was lower in this single-center study than in reports from Italy and the UK (3,4), but similar to that reported in a recent study from the US (9). The potential contributions of patient characteristics, study methodologies, and viral variants to these discrepancies merit further study. Though the mechanisms underlying RNAemia’s contributions to multi-system pathology in both the acute and post-acute phases, when persistent, remain to be elucidated, mounting evidence for its predictive value suggests that testing for SARS-CoV-2 RNAemia at presentation may help guide the triage, management, and prognosis of COVID-19.

## Data Availability

De-identified study data are presented as online datasets.

## Acknowledgements

The authors would like to thank the additional author members of the Stanford COVID-19 Biobank Study Group include: Elizabeth J Zudock, Marjan M Hashemi, Kristel C Tjandra, Jennifer A Newberry, James V Quinn, Ruth O’Hara, Euan Ashley, Rosen Mann, Anita Visweswaran, Thanmayi Ranganath, Jonasel Roque, Monali Manohar, Hena Naz Din, Komal Kumar, Kathryn Jee, Brigit Noon, Jill Anderson, Bethany Fay, Donald Schreiber, Nancy Zhao, Rosemary Vergara, Julia McKechnie, Aaron Wilk, Lauren de la Parte, Kathleen Whittle Dantzler, Maureen Ty, Nimish Kathale, Arjun Rustagi, Giovanny Martinez-Colon, Geoff Ivison, Ruoxi Pi, Maddie Lee, Rachel Brewer, Taylor Hollis, Andrea Baird, Michele Ugur, Drina Bogusch, Georgie Nahass, Kazim Haider, Kim Quyen Thi Tran, Laura Simpson, Michal Tal, Iris Chang, Evan Do, Andrea Fernandes, Allie Lee, Neera Ahuja, Theo Snow, James Krempski. We would also like to thank Hien Nguyen, Lingxia Jiang, and Paul Hung from COMBiNATi Inc. for all the material and technical support.

