## Supplement for "Association Between SARS-CoV-2 RNAemia and Post-Acute Sequelae of COVID-19"

**Table S1. Logistic model of PASC risk.**

| **Variable** | **Odds Ratio** | **95% CI** |
| --- | --- | --- |
| Age | 0.99 | 0.99 – 1.00 |
| Male | 0.89 | 0.76 – 1.03 |
| Disease severity at enrollment (reference = mild) |  |  |
| Moderate | 1.10 | 0.92 – 1.32 |
| Severe | 1.24 | 0.84 – 1.83 |
| Duration of symptoms by enrollment | 1.01 | 0.96 – 1.02 |
| **RNAemia at enrollment** | **1.31** | **1.08 – 1.59** |
| Symptomatic at enrollment | 1.28 | 0.89 – 1.83 |

n=155

Potential predictors of Post-Acute Sequelae of COVID-19 (PASC) at follow-up included: age, gender, severity of disease upon enrollment (mild = discharged from ED, moderate = hospitalized, requiring no more than oxygen by nasal cannula, and severe = hospitalized, requiring high-flow nasal cannula or mechanical ventilation), duration of symptoms upon enrollment, RNAemia at enrollment, and presence of any symptom at enrollment (anxiety, dizziness, fatigue, hair loss, palpitations, rash, insomnia, chest pain, chills, cough, decrease in sense of taste, fever, nausea/vomiting/diarrhea, headache, loss of smell, myalgia, new confusion, shortness of breath).
